# TIME-COURSE OF NEUROPSYCHOLOGICAL FUNCTIONING IN ANEURYSMAL SUBARACHNOID HAEMORRHAGE AND ITS ASSOCIATION WITH VASOSPASM

**DOI:** 10.1101/2025.03.26.25324733

**Authors:** Giorgia Abete-Fornara, Claudia Fanizzi, Elena Scagliotti, Giorgio Fiore, Valeria Conte, Fabrizio Ortolano, Tommaso Zoerle, Marco Locatelli, Giulio Andrea Bertani

## Abstract

**Background:** Aneurysmal Subarachnoid Haemorrhages (aSAH) are a severe condition often followed by vasospasm, causing several neurological and cognitive impairments in survivors. Nowadays there is still an open debate on actual impact of aSAH and vasospasm over cognitive abilities over time. This study aims at describing cognitive functioning focusing on the acute phases after bleeding and for 18 months, and to investigate the immediate and long-term effects of vasospasm.

**Methods:** seventy adult patients have been prospectively recruited and were tested at different time points: within 48/72 hours from bleeding (T1); between 7 and 10 days after bleeding (T2); five long-term follow-ups from 1 (T3) to 18 months (T7). An extensive neuropsychological evaluation was administered, including memory, attentional and executive functions, language, praxis and level of daily functional independence. Half of the sample showed radiological vasospasm.

**Results:** results show that at T1 all tests show high percentages of impairments (ranging from 38% to 100%), in particular for visual and verbal long-term memory, constructional praxis, abstract reasoning and functional independence. Many tasks gradually improve since T2, except for executive functions and visual memory which show a slower recovery. A severe diffuse impact of vasospasm emerges at T2 but, interestingly, a linear gradual recovery already since T3 emerges for almost all the investigated functions. At the last follow-ups several tests show no significant differences between patients with and without vasospasm.

**Conclusions:** despite a severe diffuse impact of bleeding and vasospasm in the acute stages, a low prevalence of cognitive and functional impairments at the chronic phase emerges. Our data may help to better understand the cognitive and autonomy trajectories of recover over time, a helpful element for clinicians to explain and reassure patients and relatives concerning real expectations of impairments and quality of life after the aSAH, and to tailor eventual rehabilitation programs.

## INTRODUCTION

Intracranial Aneurysmal Subarachnoid Haemorrhage (aSAH) is a severe kind of stroke related to aneurysm rupture associated with high rates of morbidity and mortality due to focal and diffuse brain damages and augmented intracranial pressure. Several studies report mortality percentages of 15-35% in the first 48 hours, with rates of 50% after one month, besides recent improvements in clinical management^1–3^. Such a poor prognosis also depends on subsequent mechanisms such as vasospasm and the related delayed cerebral ischemia (DCI). The former is a very common complication consisting in blood vessels contraction, with nearly 70% of aSAH patients developing it^4^, usually starting after 3 days from first bleeding and which significantly worsens clinical outcomes and prognosis^5^. DCI is a complex ischemic event known as clinical or symptomatic vasospasm, which occurs in 17% to 40% of patients with aSAH due to arterial shrinkage.

As a result, survivors often experience severe or moderate neurological and/or cognitive impairments that interfere with everyday life functioning and autonomy^6–8^. Among the most frequently reported cognitive impairments, studies describe attention and executive functions deficits, lasting also for 1-3 years after aSAH^6,7,9^. Verbal memory is another function highly impaired both in subacute and chronic stages, with significant percentages of deficits even after several years from bleeding^10–12^; moreover, memory is reported to be the slowest function to recover compared to other cognitive and neurological abilities^10^. Impaired cognition is also responsible for difficulties in returning to work and in daily autonomy, as measured by the Activities of Daily Living Scale (ADL) and Instrumental Activities of Daily Living Scale (IADL) which have been reported to indicate a general log-term impaired autonomy, probably underestimated^13,14^.

Neural mechanisms underlying cognitive impairments and their correlations with clinical and radiological aspects are still an open debate: a systematic review concerning cognition in SAH^13^ reported inconsistent findings concerning possible correlations among neuropsychological functions and clinical parameters such as oedema, site or side of bleeding or neurological status at admission. The only stable parameter is represented by the treatment method of the aneurysm, with better cognitive outcome in patients treated with coil than clipping^3,15^, and a left side bleeding, associated with worse language outcomes^16,17^.

In general, recent studies suggest that acute clinical factors concerning a SAH, such as its severity or the thickness of the subarachnoid blood, even if involved in cognitive outcome, are not its primary determinants, as they account only for low percentages of the variability^13,18,19^. Rather, subsequent mechanisms such as vasospasm, intracranial pressure or DCI, are hypothesized to primarily contribute to neuropsychological impairments, even if opposite findings have also been described^10^.

Furthermore, data available in literature differ in the timing of the neuropsychological evaluation, with a greater amount of studies dealing with long-term cognitive outcome than with acute or subacute stages.

In this view, the present study aims to prospectively investigate the time course of the neuropsychological functioning during the first 18 months after aneurysmal SAH in adult patients, with a specific interest in the first hours/days after bleeding, and to investigate the immediate and long-term effects of vasospasm.

## MATERIALS AND METHODS

STROBE guidelines for cohort studies have been followed.

### Patients recruitment

Patients with aSAH were prospectively recruited from the Intensive Care Unit (ICU) and Neurosurgery Unit at Fondazione IRCCS Ospedale Maggiore Policlinico of Milan from November 2011 until November 2016. Patients were considered eligible according to the following criteria: a) having more than 18 years of age; b) having an aSAH confirmed at the angiography or angio-TC, independently from site or side; c) being able to undergo a neuropsychological evaluation at least in one of the first two time-points scheduled (see below); d) being able to perform an MRI and aMRI during the first 48 hours from the event. A written informed consent was signed by patients if conscious and able to understand, otherwise a significant relative signed on his/her behalf. Our local ethical committee approved this project, which is performed in line with the principles of the Declaration of Helsinki and approved by the Institutional Review Board of Foundation IRCCS Ca’ Granda Ospedale Maggiore Policlinico for studies involving humans.

### Clinical and neuropsychological evaluations

Patients underwent a complete neuropsychological evaluation at several time-points: within 48/72 hours from bleeding (acute phase, T1); between 7 and 10 days after bleeding (sub-acute phase, T2); five long-term follow-ups after 1 (T3), 3 (T4), 6 (T5), 12 (T6) and 18 months (T7). The neuropsychological battery included standardized and validated tests to assess language, short-and long-term memory for visual and verbal stimuli, constructional, ideomotor and orofacial praxis, attention and executive functions. Sustained attention is also qualitatively assessed through the observation of patients’ behaviour during the exam. Parallel versions were used when available to avoid learning effect among the evaluations. Raw scores of each test were corrected for age, educational level and gender based on Italian normative data in order to calculate the adjusted scores; equivalent scores were also calculated. In addition, functional impairments in everyday life activities were assessed through Activities of Daily Living (ADL) and Instrumental ADL (IADL) scales. The entire evaluation took approximately one hour. For the complete list of the tests and questionnaires used see **Table 1**.

**Table 1:** complete list of the neuropsychological tests and questionnaires administered, divided into respective cognitive domains.

| COGNITIVE DOMAIN | TEST | FUNCTION TESTED |
| --- | --- | --- |
| Memory | Digit Span forward (Monaco et al., 2013) | Short-term verbal memory |
|  | Corsi span (Monaco et al., 2013) | Short-term visuo-spatial memory |
|  | Rey/Taylor complex figure-recall (Carlesimo et al., 2002; Casarotti et al., 2014) | Long-term visuo-spatial memory |
|  | Rey 15- word list (RAVLT) immediate and long-term recall (Carlesimo et al., 1996) | Auditory learning/Long-term verbal memory |
| Executive functions | Digit Span backward (Monaco et al., 2013) | Verbal Working Memory |
|  | Stroop Test (Caffarra et al., 2002) | Cognitive inhibition and flexibility |
|  | Weigl Test (Spinnler et al., 1987) | Abstract and categorical reasoning |
|  | Raven Coloured Progressive Matrices (Basso et al., 1987) | Non verbal logical reasoning |
|  | Clock Drawing Test (Mondini et al., 2003) | Spatial planning, semantic memory |
| Language | Phonemic Fluencies (Novelli et al., 1986) | Verbal fluency through phonemic cue |
|  | Semantic Fluencies (Zarino et al., 2013) | Verbal fluency through semantic cue |
|  | Object naming (Catricalà et al., 2013) | Language production/semantic memory |
|  | Token Test (De Renzi et al., 1978) | Language comprehension |
| Attention | Attentive Matrices (Spinnler et al., 1987) | Visuo-spatial selective attention |
| Praxis | Rey/Taylor complex figure-copy (Carlesimo et al., 2002; Casarotti et al., 2014) | Constructional Praxis |
|  | Action imitation (De Renzi, 1968) | Ideomotor praxis |
|  | Movement imitation (De Renzi, 1966) | Oro-facial praxis |
| Daily Autonomy | Activities of Daily Living (ADL) scale (Katz et al, 1963) | Level of independance |
|  | Instrumental Activities of Daily Living (IADL) scale (Lawton et al., 1969) | Level of independance in instrumental activities |

Concerning clinical evaluations, patients were evaluated according to the World Federation of Neurological Surgeons score (WFNS). Haemorrhagic severity was evaluated through the Fisher scale calculated at the first cerebral tomography (CT).

Aneurysms were excluded from blood flow through endovascular (coil) or surgical (clip) techniques within the first 24 hours. When hydrocephalus was reported, an EVD was placed through the Kocher’s point. A 3-tesla MRI and angio-MRI were performed within 48 hours (T1) and 7-10 days after (T2) the aSAH, respectively. A daily monitoring of arterial blood flow was performed through transcranial eco-doppler (TDC).

### Statistical analysis

Analyses were performed using IBM SPSS version 26 and JASP version 0.18.3. Descriptive statistics were calculated to describe the cognitive performance of the whole sample over time. T-tests for independent samples were run to study the effects of vasospasm over cognition at T2, whilst Chi-square tests analysed the differences between patients with or without vasospasm in several clinical variables. Mann-Whitney tests were used for ADL and IADL scales due to their non-normal distributions.

Two-ways mixed repeated measures ANOVAs with Bonferroni Correction for multiple comparisons were run to study the direct and interactive effects of Time (within-subjects factor) and Vasospasm (between-subjects factor) on the adjusted scores of the cognitive tests at the different time-points. Results were considered significant with a *p* value < 0.05.

## RESULTS

### Patients cohort

Seventy adult patients were enrolled, with a mean age of 54.56 years (± 11.88) and a mean educational level of 11.10 years (± 3.93), 45 (64.3%) of whom were females. In 61 patients (87.1%) the haemorrhage occurred in the anterior circulation and in almost half of the sample (33, 47.1%) it interested the anterior communicating artery. Fifty-one subjects developed no hydrocephalus. Almost all patients (65, 92.8%) received a coiling treatment. The complete list of socio-demographical and clinical data is reported **in Table 2**.

**Table 2:** complete list of socio-demographical and clinical data of the sample. SD= standard deviation; WFNS: World Federation of Neurological Surgeons score scale; ICA: Internal Carotid Artery; MCA: middle cerebral artery; PICA: posterior cerebellar inferior artery; PCA: posterior cerebral artery.

|  | POPULATION |  |  |
| --- | --- | --- | --- |
|  | Whole sample | Vasospasm yes | Vasospasm no |
| **Age (years):** mean (SD), range | 54.56 (11.88), 26-79 | 53.83 (10.36), 31-79 | 55.29 (13.34), 26-79 |
| **Education (years):** mean (SD), range | 11.10 (3.93), 3-18 | 10.97 (4.02), 3-17 | 11.23 (3.89), 3-18 |
| **Gender, N (%)**<br>Females:<br>Males: | 45 (64.3%)<br>25 (35.7%) | 23 (65.7%)<br>12 (34.3%) | 22 (62.8%)<br>13 (37.1%) |
| **WFNS, N (%)**<br>1:<br>2:<br>3:<br>4:<br>5: | 26 (37.1%)<br>17 (24.3%)<br>11 (15.7%)<br>12 (17.1%)<br>4 (5.7%) | 9 (25.7%)<br>8 (22.9%)<br>6 (17.1%)<br>8 (22.9%)<br>4 (11.4%) | 17 (48.6%)<br>9 (25.7%)<br>5 (14.3%)<br>4 (11.4%)<br>0 (0%) |
| **Fisher scale, N (%)**<br>1:<br>2:<br>3:<br>4: | 12 (17.1%)<br>10 (14.3%)<br>23 (32.9%)<br>25 (35.7%) | 5 (14.3%)<br>5 (14.3%)<br>12 (34.3%)<br>13 (37.1%) | 7 (20.0%)<br>5 (14.3%)<br>11 (31.4%)<br>12 (34.3%) |
| **Circulus, N (%)**<br>Anterior:<br>Posterior: | 61 (87.1%)<br>9 (12.9%) | 33 (94.3%)<br>2 (5.7%) | 28 (80.0%)<br>7 (20.0%) |
| **Site of bleeding, N (%)**<br>Anterior Communicating Artery:<br>ICA:<br>Posterior Communicating Artery:<br>MCA:<br>PICA:<br>Basilar Artery:<br>PCA: | 33 (47.1%)<br>11 (15.7%)<br>10 (14.3%)<br>10 (14.3%)<br>3 (4.3%)<br>2 (2.9%)<br>1 (1.4%) | 20 (57.1%)<br>3 (8.6%)<br>2 (5.7%)<br>8 (22.9%)<br>0 (0%)<br>2 (5.7%)<br>0 (0%) | 13 (37.1%)<br>8 (22.9%)<br>8 (22.9%)<br>2 (5.7%)<br>3 (8.6%)<br>2 (5.7%)<br>1 (2.9%) |
| **Surgical Treatment, N (%)**<br>Coil:<br>Clip: | 65 (92.8%)<br>5 (7.1%) | 32 (91.4%)<br>3 (8.6%) | 33 (94.3%)<br>2 (5.7%) |
| **Hydrocephalus, N (%)**<br>Yes<br>No | 19 (27.1%)<br>51 (72.9%) | 12 (34.3%)<br>23 (65.7%) | 7 (20%)<br>28 (80%) |

Radiological vasospasm at aMRI was detected in 35 patients (50%), whose characteristics are described in **Table 2**, and occurred in the anterior communicating artery for most of the sample (20 patients, 57.1%). No significant differences emerged between patients with and without vasospasm for age or sex, whilst a significant difference (*p*=0.009) was found for sites of bleeding, independently from the haemorrhage severity as calculated with the Fisher score. In particular, as shown in **Table 2**, vasospasm was more common in the Anterior Communicating and Middle Cerebral Arteries (X^2^ =16.96). Twelve patients (17.1%) developed transient or permanent neurological impairment due to vasospasm assessed radiologically. The association between clinical vasospasm and neuropsychological assessment was not performed because of the unbalanced distribution of the sample.

As **Table 3** reports, not all patients completed the neuropsychological evaluation at all seven time-points. In the acute and sub-acute phases, a few patients did not manage to sustain the cognitive effort required to be evaluated in one of the two time-points. In a few cases, patients were lost at the follow-ups due to the following reasons: patient unwillingness to undergo the exam, patient unable to reach the hospital for clinical or personal reasons, the impossibility to contact him/her.

**Table 3:**
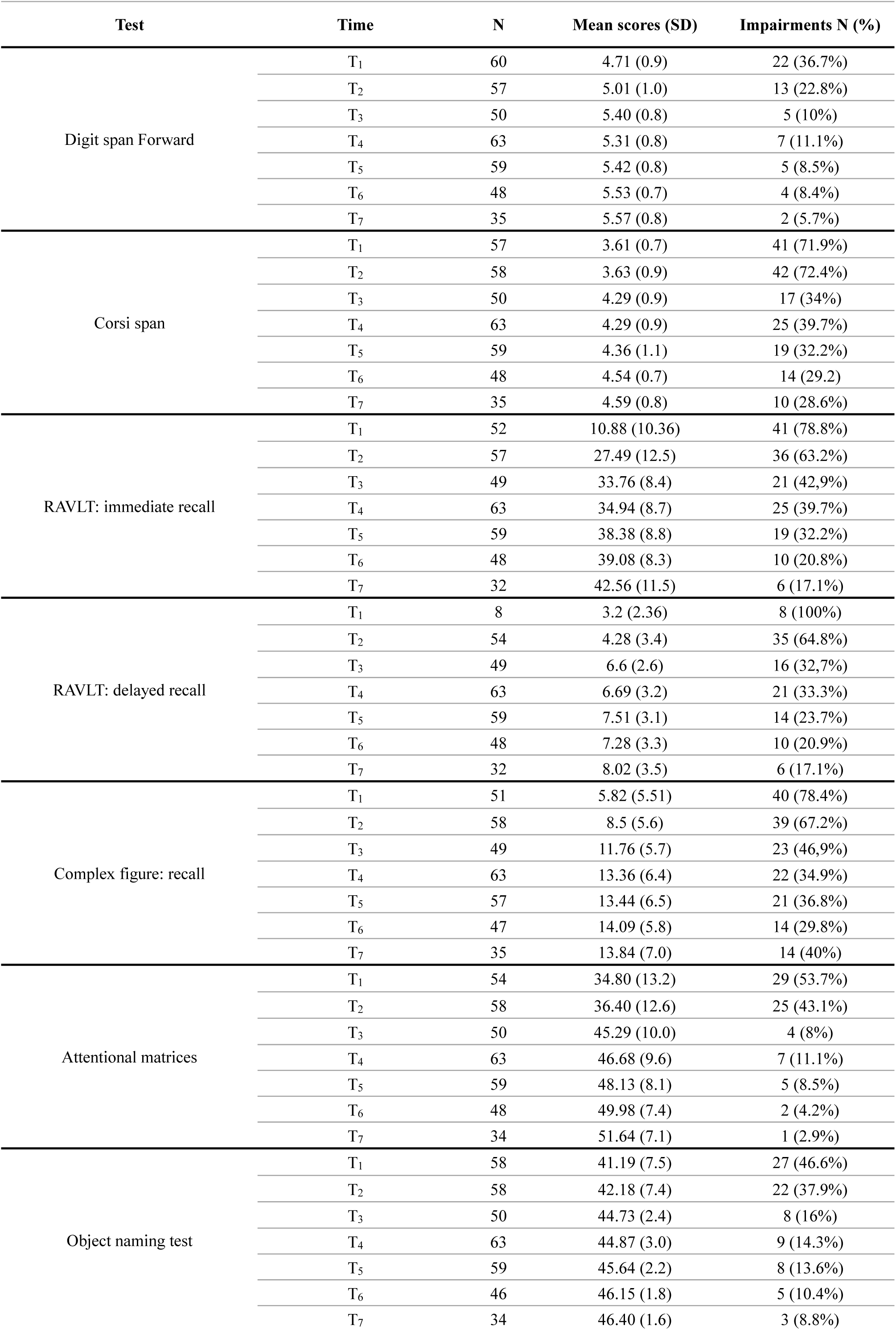
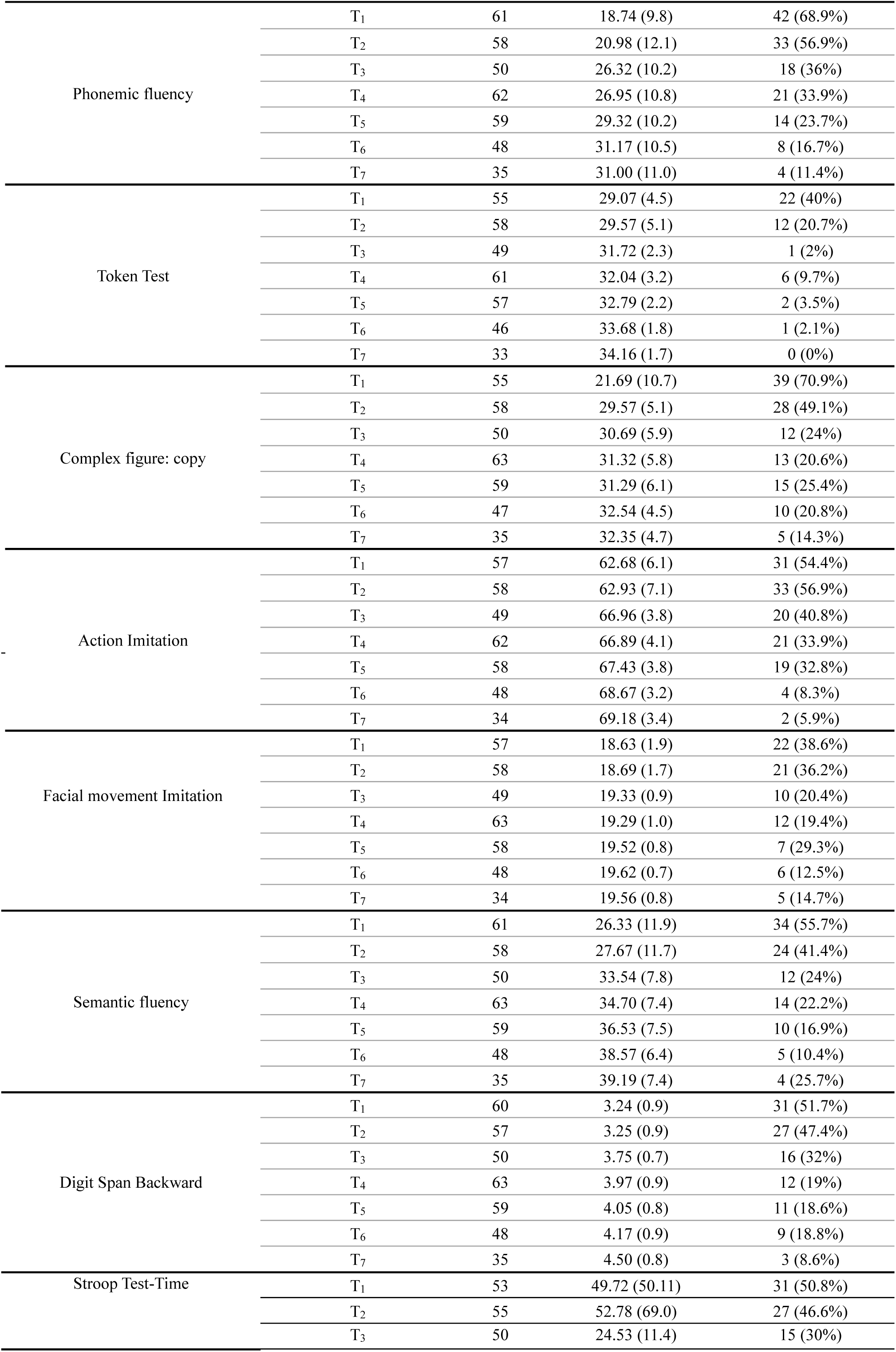
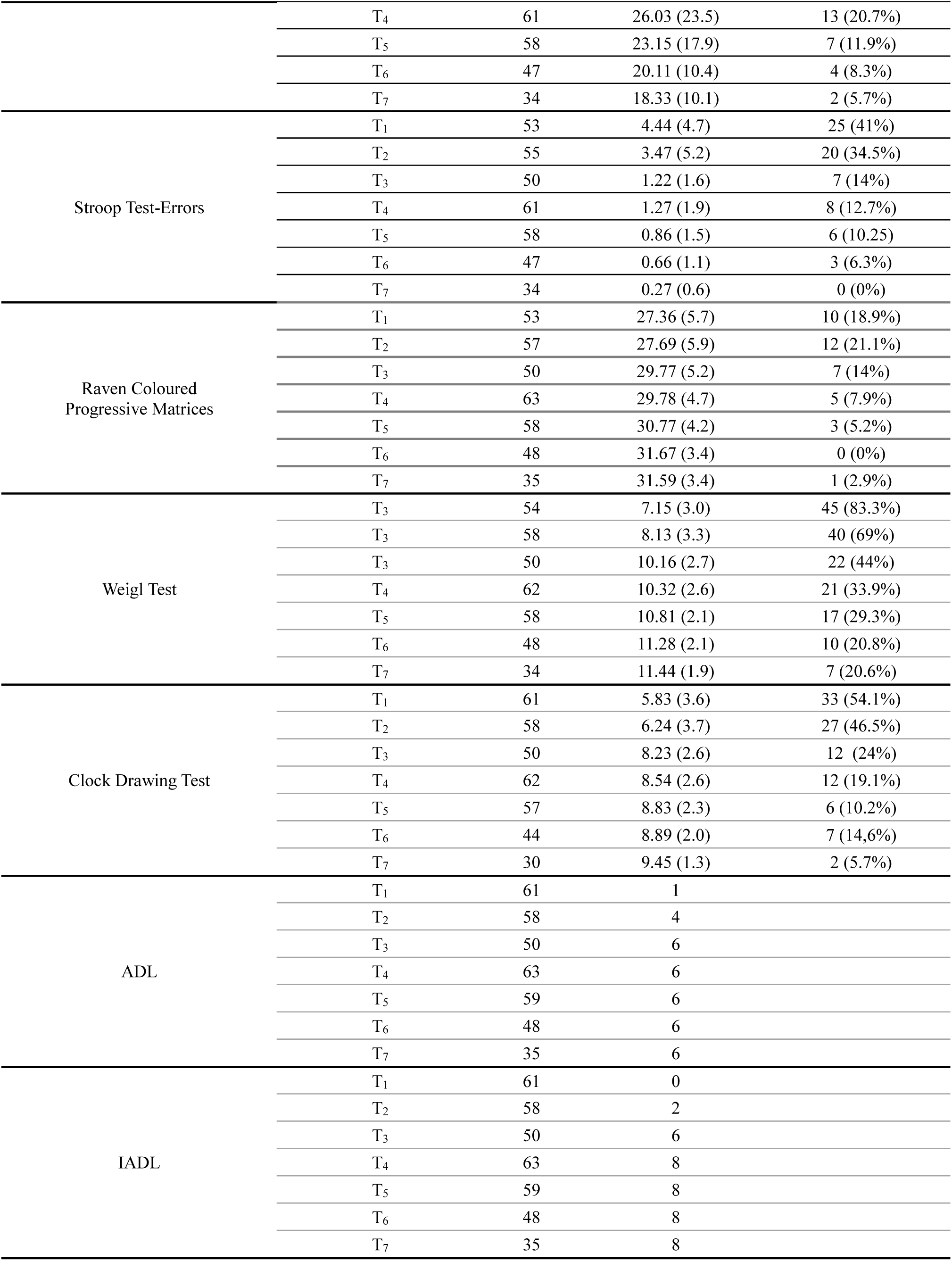
mean scores and percentages of impairments of the neuropsychological tests administered in the different time-points for the whole sample. SD= Standard Deviation.

Noteworthy, also patients with a WFNS score of 4 or 5 managed to be tested at T1 or T2 obtaining also normal scores, despite their critical clinical conditions.

### Statistical analysis

Concerning neuropsychological functioning over time, mean scores for each test and relative percentages of impairment for each time-point are reported in **Table 3**. Data show that memory is highly impaired at T1 in all the sub-domains tested, with percentages of impairment ranging from around 37 to 100%. In particular, RAVLT-Immediate and Delayed are the tests impaired the most, the latter even reports impaired functioning in the whole sample, with only 8 patients who managed to undergo it. All functions recover already in the sub-acute phase (T2), except for the Corsi span starts to improve one month after the haemorrhage and shows a gradual improvement over time. The only exception is the Figure Recall test, which maintains a 40% of impairment at T7.

Visual attention is impaired in around half (53.7%) of the sample at T1, whilst it reaches very low percentages of impairment (3%) at T7.

Concerning language, at T1 notable percentages of impairment emerge, ranging from 40 to 68.9%; however, it shows a good recovery in both production and comprehension functions over time: at T2 all tests improve, especially the Token test which halves the impaired performances, from 40% to 20.7%, and obtains a complete recovery (0%) at T7. The higher impairment at T7 concerns the Semantic Fluency (25.7%). The Phonemic Fluency is the most impaired test at T1 and shows the slowest recovery over time.

Also the praxis domains show a general improvement over time, with the Figure copy test showing the highest percentages of impairment in the acute (70.9%) and sub-acute (49%) phases.

As far as executive functions are concerned, at T1 impairments range from 20 (Raven Matrices) to 83% (Weigl test), resulting the most heterogeneous domain in the acute phase, with a mean general impairment of around 50%. Only slight recoveries appear at T2, differently from the other domains, but an evident improvement emerges during the follow-ups, with significantly lower percentages of impairments at T7, as for example the Stroop-Error score which reports absence of impaired performances. The Weigl test shows the highest impairment in this domain at the last follow-up (around 21%).

Concerning daily autonomy, a severe reduction is necessarily present at T1 in both ADL and IADL scales; interestingly, whereas ADL improves already at T2, IADL starts to improve only at T4. A complete autonomy is restored for both scales at T7.

Regarding radiological vasospasm effects on cognition at T2, the T-tests showed a significant impact on several tests (to note: no statistical differences emerge at T1 from patients with and without vasospasm): for the memory domain, patients with vasospasm had a significantly worse performance at the Corsi test (p=0.024, t_(56)_=2.32, mean score 3.34 ± 1.02) and the Figure recall test (p=0.033, t_(56)_=2.19, mean score 6.82 ± 5.85), as compared with those who did not develop it (mean scores 3.88 ± 0.71 and 9.96 ±5.08, respectively).

Patients without radiological vasospasm performed better also at the Token test (p=0.025, t_(56)_=2.45, mean 31.03 ± 3.03) and at the Phonemic fluency test (p=0.027, t_(56)_=2.27, mean 24.16 ± 11.8) with respect to those with (mean 27.89 ± 6.34 and 17.25 ± 11.48, respectively), and, concerning the executive functions, at the Clock Drawing (p=0.044, t_(56)_=2.09, mean 7.17 ± 3.35) and the Stroop Test, in both the Time (p=0.039, t_(53)_=-2.17, mean 33.45 ± 19.12) and Errors scores (p=0.032, t_(53)_=-2.26, mean 1.98 ± 2.27). Patients with vasospasm obtained a mean score of 5.16 ± 3.96 at the Clock Drawing test, whilst a mean score of 75.89 ± 96.17 was found for the Time score of the Stroop Test, showing a great difference between the two groups; their Stroop-Error mean score was 5.27 ± 6.97, instead. Similarly, the Digit Span Backward was significantly more impaired (p=0.046, t_(55)_=2.04, mean 2.45 ± 0.95) in patients with vasospasm, as compared to those without (mean 3.49 ± 0.94).

For the praxis domain, only the Figure copy test showed significant (p=0.030, t_(55)_= 2.40) lower mean scores in the vasospasm group (23.50 ± 11.67 vs 29.33 ± 6.75).

Finally, a significant impact of vasospasm was found also for the ADL (p=0.003, U=241.5) and IADL scales (p=0.004, U=241.5): patients with vasospasm were significantly more impaired in both strumental (mean 3.85 ± 2.1) and instrumental activities (mean 1.22 ± 1.08) than those who did not develop it (mean 5.29 ± 1.3, and 2.16 ± 1.21, respectively).

To further study the effects of vasospasm over time, a mixed repeated measures ANOVA with Time and Vasospasm as predictors was conducted, showing significant linear trends of improvement for Time for all the tests administered, indicating a linear gradual recovery over time for all the investigated functions. In only the Digit Span Backward a significant interaction effect (p=0.023, F_(6)_=2.56; Mauchly’s test p=0.095) between the two independent variables considered emerged. As shown in **Figure 1a**, whereas test’s scores of patients without vasospasm gradually improve over time, a swinging trend emerges for patients with vasospasm: interestingly, in both the two groups a significant difference (p=0.004 and <0.001) between respective mean scores at T1 and T7 is reported. Moreover, in the vasospasm group, significant differences emerge between T2 and all the following time-points (<0.001<p<0.038).

**Figure 1:**
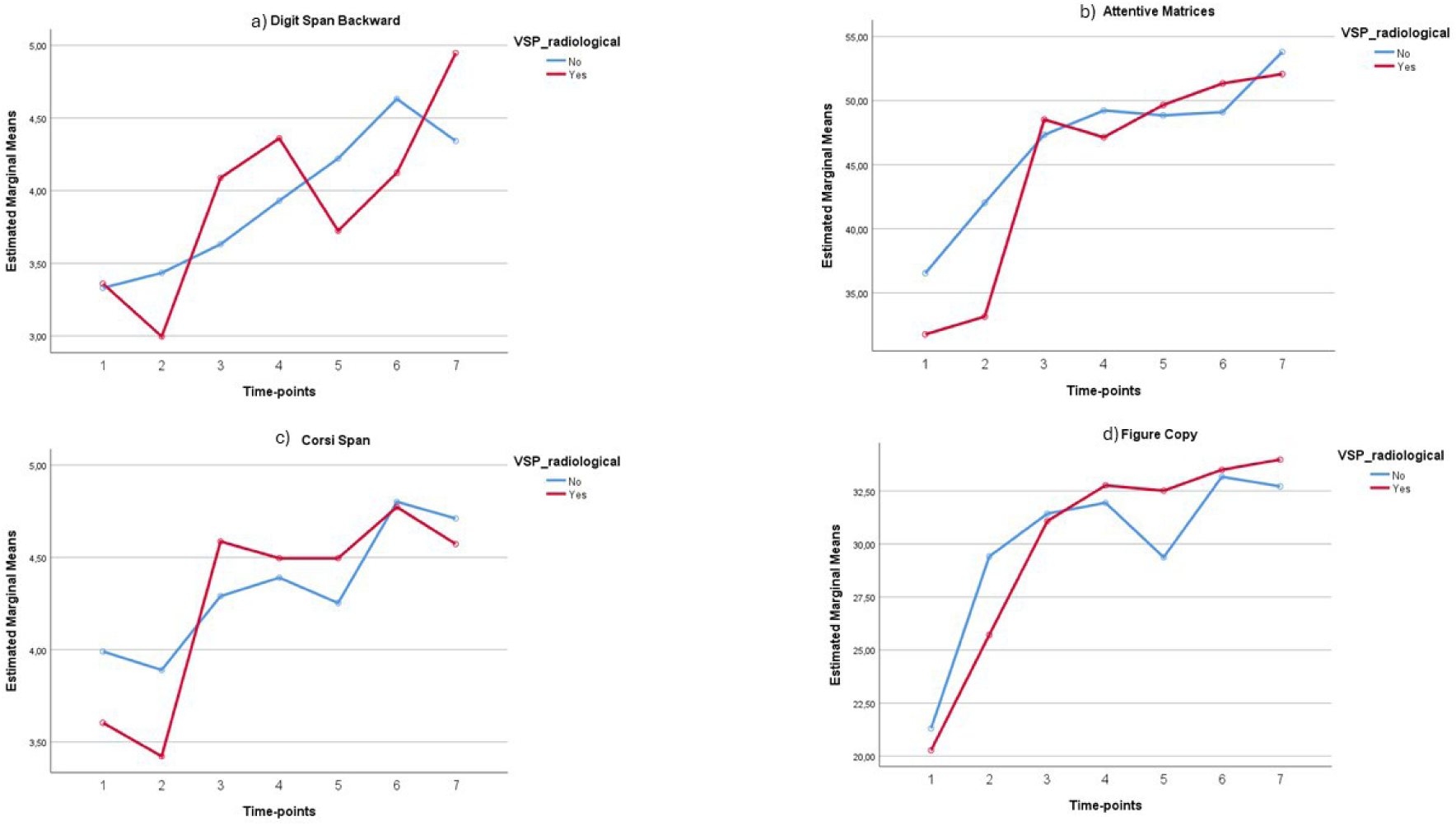
example of recover trajectories of some of the tests administered

Noteworthy, patients who experienced vasospasm show an evident improvement in most of the tests immediately at T3, often reaching at this time-point the same level of the other group, as shown in **Figure 1b-c** for an example. Only a few tests show different qualitative trends: the RAVLT Immediate and Delayed scores show an anticipated recovery at T2, and similar scores at T6 (p=1.000) and T7 (p=1.000) in the two groups indicate a general recovery of verbal learning and long-term memory independently from vasospasm. Similarly, in the Figure copy task the improvement can be observed already at T2 also for the vasospasm group (**Figure 1d**).

## DISCUSSION

In this paper we first aimed to deeply describe the neuropsychological functions over time in patients with aSAH, including levels of daily independence, focusing on the acute and sub-acute stages. In our opinion, this is of paramount importance as aSAH incidence is higher during the 40-60 years of age, a range in which people are highly productive and with significant familiar and working responsibilities. Thus, a better understanding of the cognitive and autonomy recoveries over time can be helpful for clinicians to explain and reassure patients and relatives concerning real expectations of impairments and quality of life after the aSAH, and to tailor eventual rehabilitation programs.

The cognitive and functional deficits observed in this study underscore the profound and multifaceted impact of aSAH on brain function: the acute-phase impairments, particularly in memory, attention, and executive functions, are consistent with prior studies reporting widespread cortical and subcortical damages secondary to the initial haemorrhage, elevated intracranial pressure, and delayed ischemic events. The recovery patterns across cognitive domains revealed different trajectories, with visuospatial memory and executive functions showing slower recovery compared to attention and language. This heterogeneity suggests that specific neural circuits, such as the hippocampal and prefrontal networks, may be more susceptible to ischemic and inflammatory damage, requiring longer periods for neuroplastic reorganization.

An extensive review^13^ reported a great amount of cognitive deficits during the first three months after haemorrhage, with discordant results concerning long-term outcomes. In particular, the memory domain was reported to be most compromised, whose impairments lasted also after 18 months, especially for the long-term verbal functions. Coherently, in our sample, the memory domain shows great impairments in the acute phase, markedly for verbal learning/long-term verbal memory, indicating an evident acute impact of bleeding over these functions; besides the eventual presence of vasospasm, in the sub-acute follow-up (T2) all tests improve their scores, except for the short-term visuospatial memory test (Corsi span), suggesting a slighter effect of vasospasm over these functions, as confirmed by the other analyses. Gradual improvement emerges over time, until T7 in which very low percentages of impairment were detected, in particular for the verbal short-term memory test, an element in line with previous reports^10^. Only the long-term visuospatial memory task shows a higher prevalence of deficit at the 18-month follow-up, differently from another paper in which visual memory showed complete recovery after one year from aSAH^10^.

Also, the visual attention test, despite half of the sample scoring deficiently in the acute phase, shows significant improvements over time, with only one patient reporting an impaired performance at 18 months.

In the acute phase high prevalence of impairment is visible in the language domain, in particular for the phonemic fluency test; interestingly, already at T2 a general recovery can be found, and, at 18 months, almost all patients show normal performances in the language tests. A similar pattern emerges for the praxis functions, whereas the executive functions domain does not improve in the sub-acute phase, suggesting a greater impact of vasospasm over this kind of cognitive abilities. Nevertheless, also the executive domain gradually improves over time, with very low percentages of impairment at the chronic stages. A greater variability of trends and impairments has been described^13^ for executive functions, with respect to the other cognitive domains which show more stable and gradual trends of improvement over time^12^.

For daily autonomy, severe impairments are present in the acute phase, coherently with previous data^13^, confirming the great impact of aSAH on functional status. Noteworthy, a complete recovery can be depicted at the chronic stages.

In our sample, patients with vasospasm obtained significantly worse scores at T2 for long- and short-term visual memory, constructional apraxia, language comprehension, phonemic fluencies, verbal working memory, cognitive inhibition and daily independence. Thus, vasospasm had a deep impact on the general cognitive functioning, and in particular in visuospatial abilities, as suggested by Corsi span, Complex figure copy and recall and the Clock drawing tests. Nevertheless, while it exacerbates severe acute cognitive impairments, its long-term effects appear limited. The transient impact of vasospasm aligns with emerging evidence suggesting that global mechanisms such as neuroinflammation, rather than localized ischemia, may play a more critical role in determining chronic outcomes^20,21^. Nonetheless, the unstable recovery trends observed in working memory for patients with vasospasm raise questions about potential delayed or subtle effects of vasospasm on specific cognitive functions. Interestingly, these findings suggest that a timely management of vasospasm may mitigate its acute impact, supporting the importance of interventions like hemodynamic therapy, nimodipine administration, and close monitoring with transcranial Doppler. Moreover, the rapid recovery observed in some domains post-vasospasm highlights the resilience of certain cognitive networks, which may benefit from targeted treatments and rehabilitation during this critical period.

These findings have significant implications for patient counselling and rehabilitation planning. For instance, clinicians can reassure patients and caregivers about the expected timeline of functional recovery while emphasizing the importance of continued cognitive and physical therapy.

The study indirectly supports the hypothesis that mechanisms such as neuroinflammation, diffuse axonal injury, and blood-brain barrier disruption may underlie the observed cognitive impairments and recovery patterns. Previous research has linked elevated levels of inflammatory biomarkers (e.g., interleukin-6 and C-reactive protein) with worse cognitive outcomes following aSAH^21,22^. Understanding these mechanisms could pave the way for novel therapeutic approaches, such as anti-inflammatory treatments or neuroprotective agents, to enhance recovery.

In this view, incorporating regular neuropsychological assessments into standard care protocols could also help identify patients at risk of prolonged deficits and guide timely interventions.

In general, in our sample, the variable Time seems to be more efficient in predicting cognitive outcomes than the presence of vasospasm: despite being a severe condition, patients who survive an aSAH and the first treatments show a low prevalence of cognitive impairment at the chronic stages and no impairments in daily autonomy. Even patients with vasospasm report slight long-term effects of this event, a reassuring element to take into account for clinicians. Additionally, these data underline the security and efficacy of the modern treatments available for this condition, also in the acute phases.

## LIMITS AND FUTURE DIRECTIONS

A first methodologic limit is represented by the reduction in sample size over time, which limited the statistical power of the longitudinal analyses. Future prospective studies might include a fixed number of patients for all the time-points to study in order to obtain more powerful statistical data concerning long-term follow-ups. Another limit is represented by the unequal distribution of some other clinical factors (for example, hydrocephalus, site of bleeding etc) limiting the exploration of their effects and interactions. Future studies with stratified recruitment or larger sample sizes could address this issue. Moreover, in future research, the inclusion of advanced neuroimaging techniques, such as diffusion tensor imaging (DTI) or resting-state functional MRI, could provide deeper insights into structural and functional changes associated with recovery.

## SOURCES OF FUNDING

none

## DISCLOSURES

none

## Data Availability

All data are available in the Institution datasets by asking the corresponding author

## Notes

### Competing Interest Statement

The authors have declared no competing interest.

### Funding Statement

No funding have been received

### Author Declarations

Our local ethical committee approved this project, which is performed in line with the principles of the Declaration of Helsinki and approved by the Institutional Review Board of Foundation IRCCS Ca' Granda Ospedale Maggiore Policlinico for studies involving humans.

